# Inconsistency of infant caretakers’ visual exposome with safe infant sleep recommendations: a systematic review and meta-analysis

**DOI:** 10.1101/2024.10.14.24315451

**Authors:** Sophie de Visme, Yaël Pinhas, Jérémie F. Cohen, Rachel Y. Moon, Floortje Kanits, Sabine Plancoulaine, Anne-Laure Sellier, Inge Harrewijn, Christèle Gras-Le Guen, Martin Chalumeau

**Author notes:** Corresponding author Sophie de Visme, Nantes Clinical Investigation Centre, Nantes University Hospital, 5 allée de l’île gloriette, 44093 Nantes CEDEX 01, France. E-mail address.

## Abstract

**Importance:** Rates of sudden unexpected death in infancy (SUDI) remain high in several high-income countries. Several studies reported rates of pictures of sleeping infants or infant sleeping environments that were highly inconsistent with safe infant sleep recommendations (SISRs) to prevent SUDI.

**Objective:** To provide robust and synthetized evidence to decision-makers involved in corrective actions, we systematically assessed the proportion of pictures that were inconsistent with SISRs in the visual exposome of infant caretakers.

**Data Sources:** In November 2023, we searched PubMed, Web of Science, and Google Scholar.

**Study Selection:** Eligible studies included were those reporting the quantitative evaluation of the inconsistency between one or several SISRs and pictures depicting sleeping infants or a sleeping environment intended for an infant in physical and digital public spaces.

**Data Extraction and Synthesis:** Data were extracted independently by 2 authors. The risk of bias of included studies was assessed with a customized version of Hoy’s tool for prevalence studies. A random-effects logistic regression model was used to obtain summary estimates of proportions of pictures. Between-study heterogeneity was estimated with the *I*^2^ statistic.

**Main Outcomes(s) and Measure(s):** The proportion of pictures inconsistent with one or several SISRs from the American Academy of Pediatrics.

**Results:** We screened 1,086 articles and included 7 studies conducted between 2008 and 2023 that analyzed pictures found in parenting magazines, online and print newspapers, baby diaper packaging, commercial stock photography websites, and Instagram. The overall risk of bias was deemed low. Among the 5,442 pictures depicting sleeping infants or infant sleep environments, the summary estimates of the proportion of inconsistencies with SISRs were 39% for a non-supine sleeping position, 5% for a soft sleeping surface, 8% for sharing the sleeping surface, 14% for an unsafe crib, 58% for soft objects or loose bedding, 17% for a covered head, and 85% for at least one SISR inconsistency. All summary estimates had significant between-study heterogeneity.

**Conclusions and Relevance:** Infant caretakers’ visual exposome is greatly inconsistent with SISRs and could lead to dangerous practices, which should prompt actions from manufacturers, advertisers, newspaper and website editors, social media moderators, and legislators.

## INTRODUCTION

Sudden infant death syndrome (SIDS) and accidental suffocation and strangulation in bed are the most common causes of death found after investigation of sudden unexpected infant death (SUDI).^1–4^ In the 1990s, identified risk factors for SIDS were a non-supine sleeping position and other factors related to the sleeping environment, including a non-firm sleep surface, lack of room sharing with parents, sharing the sleep surface with another person, a non-safe crib, soft objects or loose bedding, and head covering.^5–11^ Although the prevention campaigns of the 1990s, consisting of safe infant sleep recommendations (SISRs), succeeded in reducing the incidence of SIDS by 50% to 80% depending on the country, since the early 2000s, the incidence has decreased only slightly or has even plateaued.^12–14^ In recent studies conducted in Australia, France, The Netherlands, and the United States, rates of parental practices for infant sleep that were inconsistent with SISRs ranged from 19% to 34%.^15–19^

Among the various factors reported to influence parents’ and, more generally, infant caretakers’ behaviors,^20^ images act via the influence mechanisms of authority, social proof, and unity.^21–23^ Images are well established as a modality better remembered than words, a phenomenon known as the “picture-superiority effect”.^24–27^ The potential power of images to affect human health behavior led to their use in campaigns to prevent smoking,^28–31^ alcohol consumption,^32^ sun exposure,^33^ and obesity.^34,35^ Their effectiveness in influencing human behavior has led to legislation by some public authorities, for example, by the mandatory printing of a pictogram discouraging alcohol consumption during pregnancy on all alcohol bottles^36^ or by banning images of babies on infant formula packaging in Europe to avoid promoting artificial feeding to the detriment of breastfeeding.^37^

Several studies have evaluated the level of inconsistency between SISRs and pictures of sleeping infants or sleeping environments intended for an infant in the physical or digital visual public spaces. These studies reported variable but sometimes high rates of inconsistency with SISRs.^38–40^ Together, these studies provide an overview of what can be referred to as “the visual exposome” of infant caretakers, that is, the sum of pictures to which infant caretakers are exposed and that may influence their knowledge, attitudes, and practices regarding the implementation of SISRs. Of concern, these publications have not yet resulted in any corrective action by private or public stakeholders. To provide robust and synthetized evidence to public and private decision-makers who are involved in such corrective action, we systematically assessed the proportion of pictures of sleeping infants or infant sleep environments that were inconsistent with SISRs in the visual exposome of infant caretakers.

## METHODS

This systematic review aimed to identify and synthesize studies evaluating the level of inconsistency between SISRs and pictures depicting sleeping infants or infant sleep environments in physical and digital visual public spaces. We followed the Center for Reviews and Dissemination’s guidance and the Preferred Reporting Items for Systematic review and Meta-analysis studies statement (PRISMA) to perform and report this study, respectively (**eTable 1**).^41,42^

### Information sources, search strategy, selection criteria and data collection process

In November 2023, we searched PubMed and Web of Science electronic databases, combining the following keywords in English language only: infant, baby, sleep, sleeping, safe, safety, unsafe, environment, recommendation, guideline, and guidance (**eText**). Two authors (YP and SV) independently assessed results by screening titles and abstracts, then full texts, to include relevant studies. In case of disagreement, consensus was reached with the help of a third author (MC). For all included studies, we examined their reference lists, searched the Science Citation Index and Google Scholar for studies citing them, and examined their first 50 related articles in PubMed.

Eligible studies were those reporting the quantitative evaluation of the inconsistency between one or several SISRs by the American Academy of Pediatrics (AAP) (i.e., supine sleeping position, a firm sleep surface, room sharing with parents, not sharing the sleep surface with another person, use of a safety crib, no soft objects or loose bedding, and no head covering)^43,44^ and pictures depicting sleeping infants or a sleeping environment intended for an infant in the physical and digital visual public spaces (e.g., magazines, newspapers, billboards, advertising leaflet, packaging of childcare products as well as television, movies, social networks, websites). We chose not to study the use of a pacifier because this SISR is not mentioned in several European recommendations and is debated.^45–47^

Two authors (SV and YP) independently extracted data on the characteristics of studies (design, setting, dates, countries), information needed for the risk of bias assessment, and numbers needed for the meta-analysis.. When data were missing or unclear in a study, the corresponding authors of the study were contacted for additional information.

### Risk of bias assessment

The risk of bias of included studies was assessed with a customized version of Hoy’s tool for prevalence studies.^48^ Customization was required because Hoy’s tool was originally developed for human participants rather than pictures in a non-finite space (**see eTable 2**). Seven of the 10 items of Hoy’s tool were retained and adapted to our study question: 5 related to internal validity (data directly collected, acceptable case definition, reliable and valid evaluation, same mode of data collection, and appropriate numerator[s] and denominator[s] for the parameter of interest) and 2 to external validity (representativeness of the target “population” and random selection). Two authors (SV and MC) independently assessed each risk-of-bias item. For each item and each study, a binary score was assigned: 0 for low risk and 1 for high risk. If an item had an unclear assessment, it was scored as high risk. As suggested,^49^ the 7 unit scores were summed to obtain an overall score classified into 3 levels of risk of bias: low = 0-2; moderate = 3-5; and high = 6-7.

### Statistical analysis

For the included studies, we described the settings and dates, the explored components of the SISRs and visual exposome, the risk of bias, and the results regarding the proportion of pictures inconsistent with SISRs. We performed a meta-analysis when 2 or more studies reported picture proportions for the same SISR, using a random-effects logistic regression model to obtain summary estimates of proportions and their 95% confidence intervals (CIs), as recommended.^50,51^ Forest plots were used to display the results. Between-study heterogeneity was estimated with the *I^2^* statistic^52^ and interpreted as per the Cochrane collaboration guidance.^52^ Statistical analyses involved using R 4.1.2 (R Core Team 2021. R: A language and environment for statistical computing. R Foundation for Statistical Computing, Vienna, Austria. URL https://www.R-project.org/) with the *meta* package (v5.5-0; Balduzzi, Rücker & Schwarzer, 2019).^53^

## RESULTS

### Study characteristics

Among the 1,086 unique studies identified by our search strategy, 8 met our inclusion criteria, but one had to be excluded because the published results did not allow for a meta-analysis and the corresponding author did not respond to requests for additional information.^54^ The 7 included studies (**Figure 1**)^40,55–60^ were conducted from 2008 to 2023 in the United States,^56,58–60^ the United Kingdom,^57^ The Netherlands,^40^ and 11 European countries^55^ (**Table 1**).

**Figure 1.**
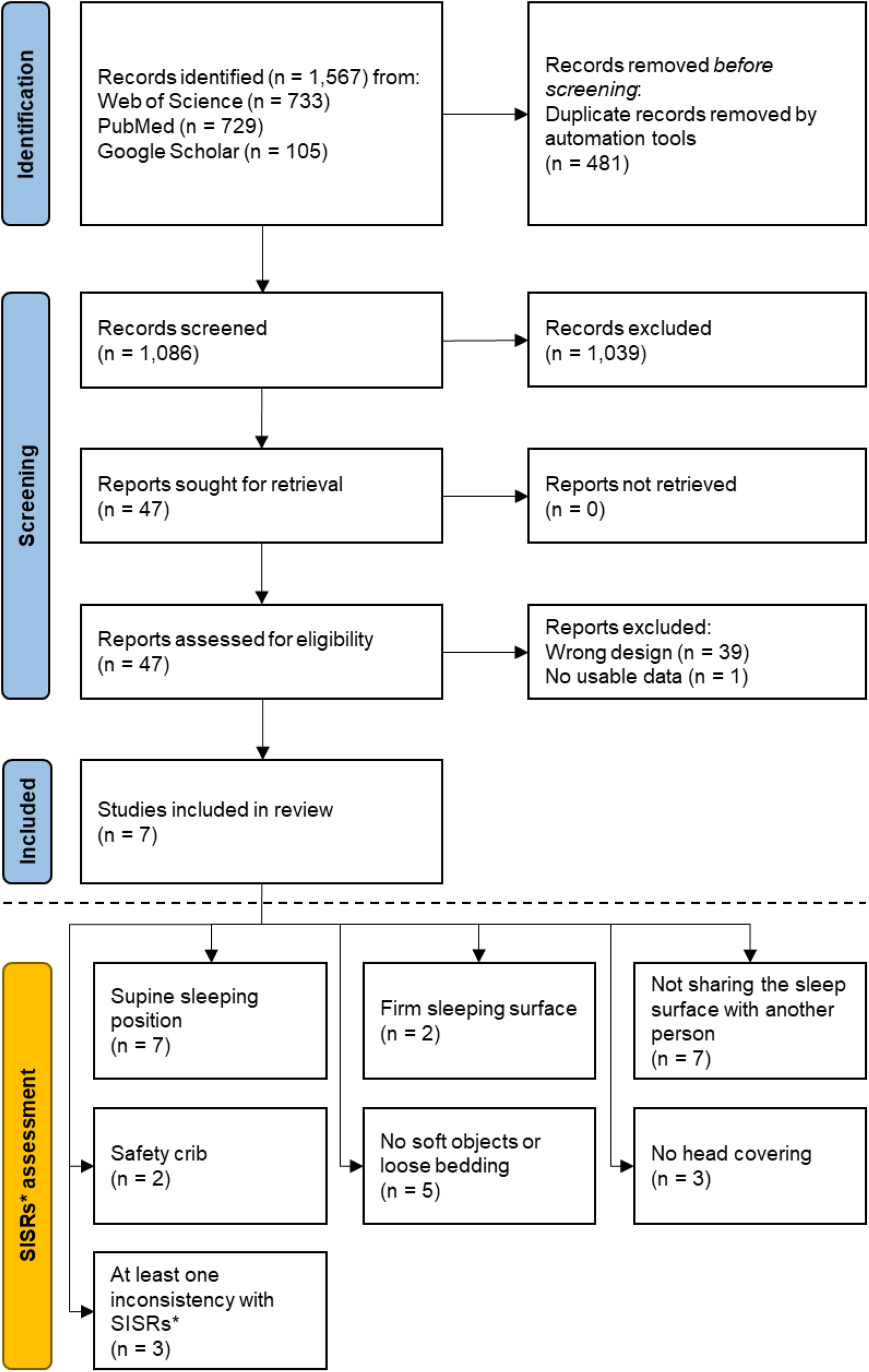
PRISMA flow diagram of the systematic review.

**Table 1.** Characteristics of included studies.

| Author<br>(year) by date<br>of publication | Country | Subject of<br>study | Study<br>period | Number<br>of pictures<br>evaluated | Number of<br>pictures<br>depicting a<br>sleeping<br>environment<br>intended for an<br>infant | Number of<br>pictures with a<br>sleeping infant | Safe infant sleep recommendations (SISRs) assessed |  |  |  |  |  |  |
| --- | --- | --- | --- | --- | --- | --- | --- | --- | --- | --- | --- | --- | --- |
|  |  |  |  |  |  |  | Supine<br>sleeping<br>position | Firm<br>sleeping<br>surface | Not<br>sharing<br>the<br>sleep<br>surface<br>with<br>another<br>person | Safety<br>crib | No soft<br>objects<br>or loose<br>bedding | No head<br>covering | At least<br>one<br>inconsist<br>ency<br>with<br>SISRs |
| Joyner *et al*<br>(2009) | USA | Printed<br>magazines | 2008 | 526 | 391 | 122 | ✓ | ✓ | ✓ | ✓ |  |  |  |
| Epstein *et al*<br>(2011) | UK | Printed<br>magazines<br>and Internet | 2009-11 | 559 | 559 | 559 | ✓ | ✓ | ✓ |  |  | ✓ |  |
| Goodstein *et al*<br>(2018) | USA | Stock photo<br>websites | 2017 | 1,947 | 1,867 | 1,590 | ✓ |  | ✓ |  | ✓ | ✓ | ✓ |
| Kamra *et al*<br>(2018) | USA | Online and<br>print<br>newspapers | 2016 | 41 | 39 | 22 | ✓ |  | ✓ |  | ✓ |  | ✓ |
| Chin *et al*<br>(2021) | USA | Instagram | 2020 | 5,400 | 1,563 | 1,134 | ✓ |  | ✓ | ✓ | ✓ | ✓ |  |
| Kanits *et al*<br>(2023) | The<br>Netherlands | Instagram | 2022 | 514 | 514 | 514 | ✓ |  | ✓ |  | ✓ |  |  |
| de Visme *et al*<br>(2023) | 11 European<br>countries | Internet | 2022-23 | 631 | 209 | 311 | ✓ |  |  |  | ✓ |  | ✓ |
| Total (number<br>of pictures,<br>studies) |  |  |  | 9,618 | 5,442 | 4,252 | 7 | 2 | 6 | 2 | 5 | 3 | 3 |

The 7 studies analyzed 9,618 pictures: 526 from print magazines “targeted to women of childbearing age”,^56^ 559 from UK “popular” magazines or “UK sleeping baby images” found with a Google search,^57^ 1,947 from the top 3 stock photo websites found with a Google search,^58^ 41 from Google News archives of online and print newspapers that covered the 2016 new AAP sleep guidelines,^59^ 5,400 from Instagram using hashtags relevant to infant sleeping practices and environments,^60^ 514 from Instagram using hashtags relevant to infant sleeping practices and from accounts of Dutch companies or platforms related to infants,^40^ and 631 from a Google search for baby diaper packages for sale online (**Table 1**).^55^ Two studies included only pictures with a sleeping infant.^40,57^ Overall, 5,442 pictures depicted a sleeping environment intended for an infant, including 4,252 pictures with a sleeping infant (**Table 1**).

The 7 included studies reported data on the following SISRs (**Table 1**): supine sleeping position (n = 7 studies; 2,100 pictures), firm sleeping surface (n = 2 studies; 29 pictures), not sharing the sleep surface with another person (n = 7 studies; 688 pictures), safety crib (n = 2 studies; 494 pictures), no soft objects or loose bedding (n=5 studies; 2,901 pictures), no head covering (n = 3 studies; 554 pictures), and at least one inconsistency with SISRs (n = 3 studies; 1,769 pictures). No study evaluated the SISR related to room sharing with parents.

### Risk of bias of included studies

With the modified Hoy’s tool, the overall risk of bias was deemed low for the 7 included studies (**eTable 3**).^40,55–60^ For the internal validity criteria, one study was judged at high risk of bias regarding the method of data collection used, which was not the same for all pictures.^57^ For the external validity criteria, all studies were judged at high risk of bias regarding the representativeness of the “target space”.

### Level of inconsistency of pictures with SISRs

Among the pictures depicting sleeping infants, a summary estimated proportion of 39% (95% CI 25-56) showed infants in a non-supine sleeping position (**Figure 2A**), 5% (95% CI 2-16) infants sleeping on a soft surface (**Figure 2B**), and 8% (95% CI 4-16) infants sharing the sleep surface with another person (**Figure 2C**). Among the pictures depicting sleeping environments intended for an infant, with or without a sleeping infant, a summary estimated proportion of 14% (95% CI 4-39) showed an unsafe crib (**Figure 3A**), 58% (95% CI 38-76) included the presence of soft objects or loose bedding (**Figure 3B**), and 17% (95% CI 15-20) showed the infant’s head covered (**Figure 3C**); 85% (95% CI 66-94) had at least one inconsistency with SISRs (**Figure 3D**). All summary estimates had a considerable between-study heterogeneity (*I^2^*>99%, p < 0.01).

**Figure 2.**
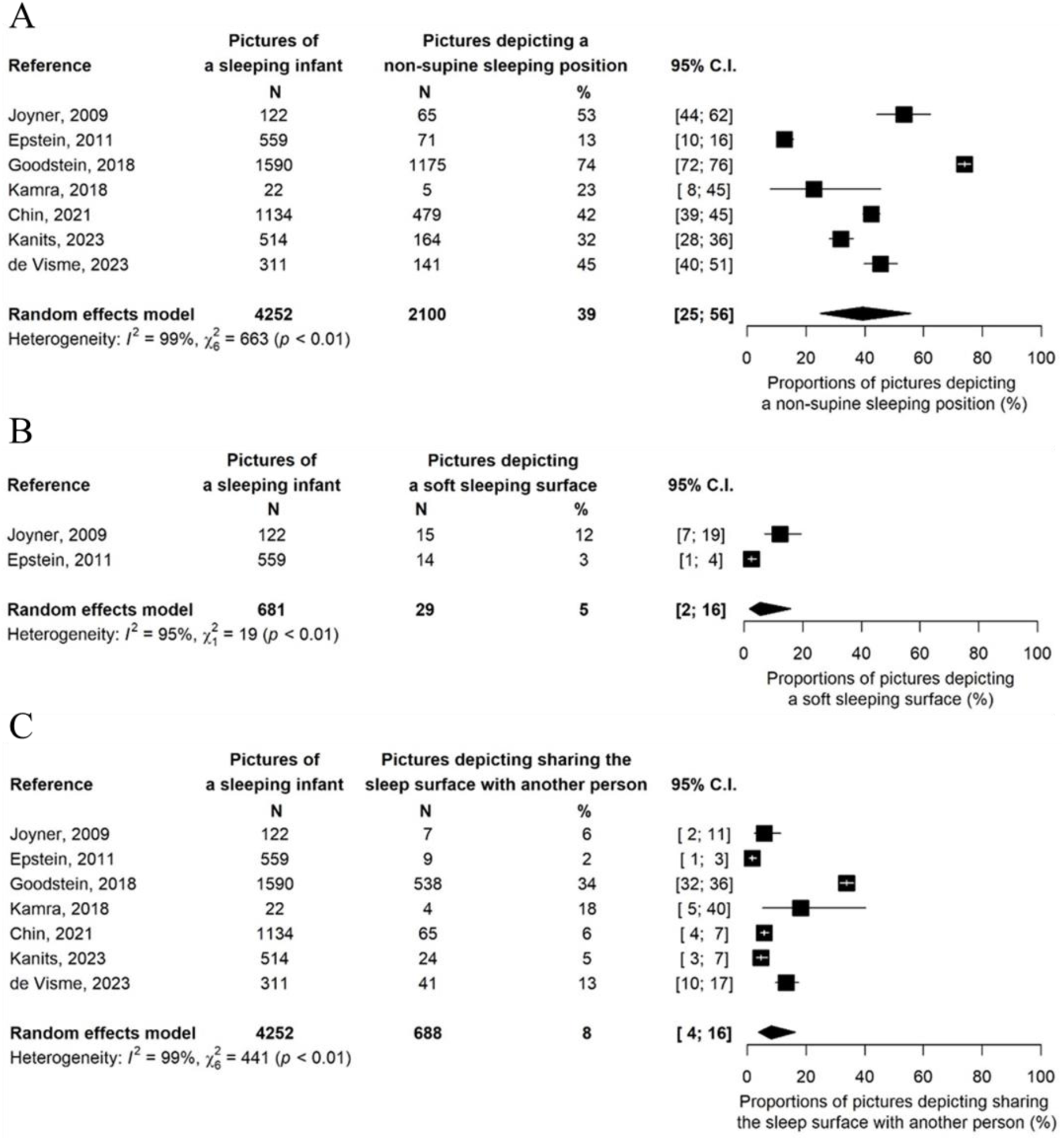
Forest plots of proportions of pictures depicting (A) a sleeping infant in a non-supine sleeping position, (B) a sleeping infant on a soft surface, and (C) a sleeping infant sharing the sleep surface with another person.

**Figure 3.**
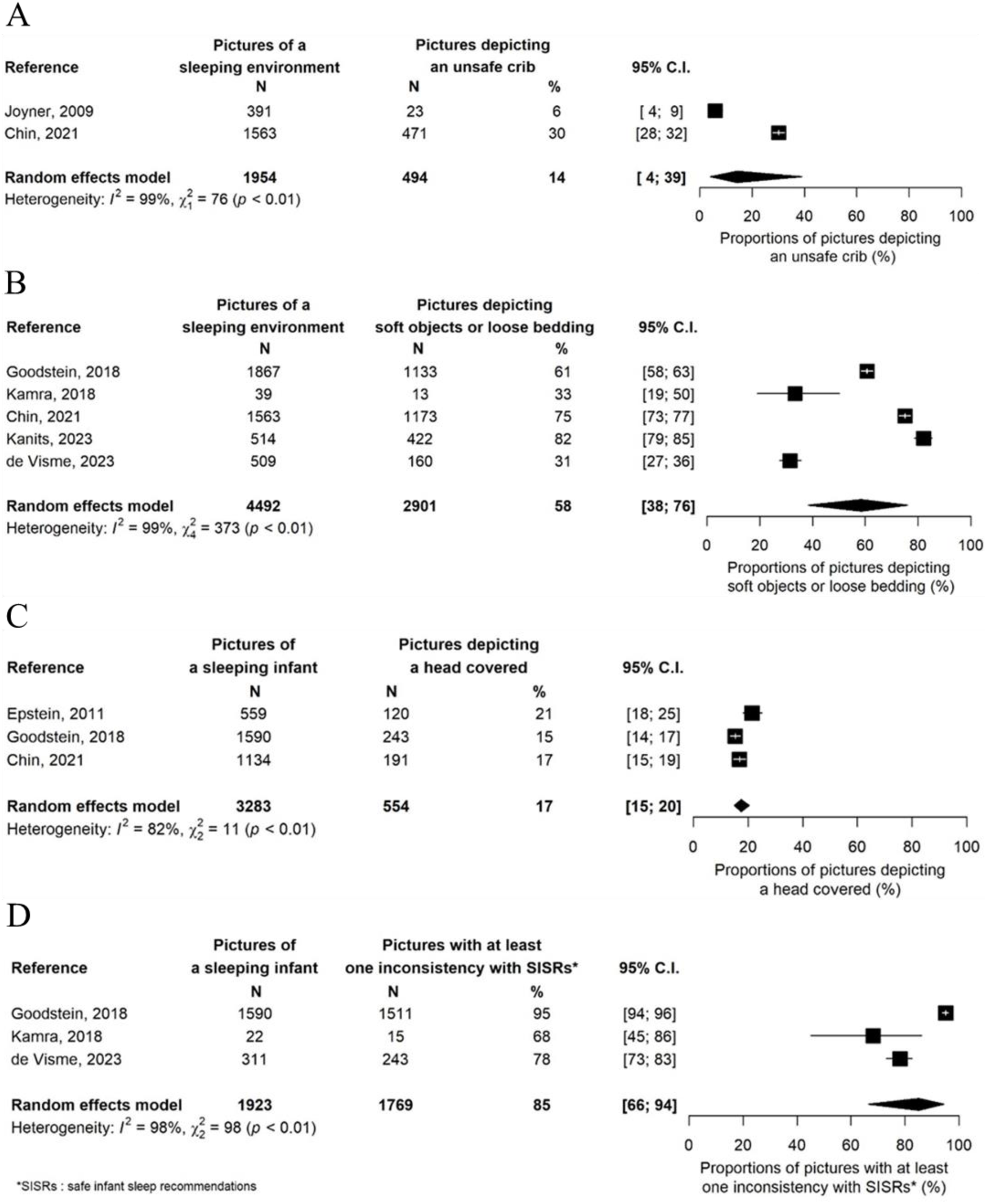
Forest plots of proportions of pictures depicting (A) a sleeping infant in an unsafe crib, (B) a sleeping environment with or without an infant, depicting soft objects or loose bedding, (C) a sleeping infant with the head covered, and (D) a sleeping infant with at least one inconsistency with SISRs.

## DISCUSSION

### Main results

In this first systematic review and meta-analysis of the proportion of pictures depicting a sleeping environment intended for an infant, with or without a sleeping infant, in the physical and digital visual public spaces, the summary estimate of the proportion of pictures that were inconsistent with at least one SISR was 85%. These pictures depicted infants in environments that have been shown in epidemiological studies to greatly increase SIDS risk. In particular, 39% of pictures were inconsistent with the back-to-sleep recommendation, with the odds of SIDS being 4.9 times increased (95% CI 3.6–6.6)^61^ when a non-supine position is used and the back-to-sleep campaigns being the major drivers for the decrease in SIDS incidence in the 1990s and early 2000s. The other inconsistencies we assessed had picture proportion estimates of 5% to 58% and portrayed environments that are well established to increase the risk of SIDS: the use of a non-firm sleeping surface, sharing a sleep surface with another person,^62–64^ using a non-safe crib, using soft objects or loose bedding, and head covering. Although room-sharing has long been recognized as a protective factor against SIDS, with a 20% to 50% risk reduction,^65,66^ we found no study that analyzed it, probably because this recommendation is difficult to assess from pictures mainly focusing on the infant and the infant’s sleep surface.

### Limitations

The first limitation is that the search strategy used only 3 electronic databases and English keywords, and some studies may have been missed, but we tried to minimize their numbers by searching Google Scholar, a multi-database search engine, for studies citing included studies. Second, for reasons described previously, we could not include one eligible study by Kreth *et al.*, who analyzed the level of inconsistency of printed advertisements from parenting magazines with the SISRs from the AAP.^54^ In this study, performed in 2015, 35% of the 428 pictures analyzed depicted infant sleep environments inconsistent with SISRs, in line with the results of the present meta-analysis. Third, included studies more often analyzed the digital (n = 7) than physical (n = 3) visual exposome, probably because digital studies are easier to conduct. Furthermore, the included studies did not analyze several static media such as specialized newspapers or magazines, billboards, or posters on public transportation and videos in television, movie theaters, or the Internet (especially social networks). Fourth, although the 7 included studies were considered to have low overall risk of bias, the representativeness of the target visual spaces studied was evaluated at high risk of bias for all 7 studies. The adaptation of Hoy’s tool is also a limitation but was necessary given the non-finite nature of the visual exposome. Fifth, for some SISRs (e.g. a firm sleeping surface) the number of inconsistencies was small, which resulted in a large confidence interval of the summary estimate. Finally, we found considerable between-study heterogeneity, which can be explained by the small number of studies included as compared with the large number of potential sources of variation including the countries studied, the study periods, and the variability of visual public spaces evaluated. It can also be explained by the partial unsuitability of use of the *I^2^* statistic to interpret in prevalence meta-analyses because it does not provide direct information about the distribution of effects.^67^ The strength and direction of the bias related to these limitations are difficult to anticipate, and the estimates provided by the meta-analyses should be considered as general indicators rather than a precise evaluation of the entire visual exposome of infant caretakers.

### Implications

Worldwide, previous SIDS prevention campaigns mainly focused on informing infant caretakers about SISRs. In one of the studies included in the present systematic review, Kamra *et al.* showed that 23% of pictures associated with the media coverage of the 2016 SISRs from the AAP depicted sleeping infants in a non-supine sleeping position.^59^ This is an issue because pictures that are not aligned with the 2016 SISRs can trigger an illusory truth effect, the finding that individuals tend to believe repeatedly presented false information as true.^68,69^ Thus, academic actors must now need to understand that producing evidence-based SISRs is not sufficient to deliver an adequate message to infant caretakers. Academic actors should embrace their societal responsibility to advocate for a safe visual exposome that conveys messages that promote SISRs and positively modify infant caretakers’ behaviors. Recently, several major US retailers have stopped selling weighted infant sleep products that are inconsistent with SISRs.^70^ Along with the results of our systematic review, these concrete actions are an example of potential future steps that public health decision-makers, manufacturers, advertisers, newspaper and website editors, social media moderators, and legislators can take in terms of internal policies, quality standards, and legislation. As has been demonstrated by successful coordinated campaigns to decrease tobacco and alcohol consumption, these type of interventions will optimize the visual exposome of infant caretakers and help to spread behavior consistent with SISRs.

## Conclusion

In this systematic review and meta-analysis, the infant caretaker visual exposome was greatly inconsistent with SISRs. Internal policies, quality standards and legislation can be used to create pictures of sleeping environments intended for an infant, with or without depicting a sleeping infant, in the physical and digital visual public spaces that are consistent with SISRs. The prevention of SUDI requires action from manufacturers, advertisers, newspaper and website editors, social media moderators, and legislators to minimize infant caretakers’ high exposure to pictures that may lead to dangerous practices and infant deaths.

## AUTHOR STATEMENTS

### Data statement

Sophie de Visme had full access to all the data in the study and takes responsibility for the integrity of the data and the accuracy of the data analysis.

### Author’s contributions

Design of the study: Sophie de Visme and Martin Chalumeau Data collection: Yaël Pinhas and Sophie de Visme

Data analyses: Sophie de Visme, Yaël Pinhas, Jérémie Cohen and Martin Chalumeau Drafting of the manuscript: Sophie de Visme and Martin Chalumeau

Revising the protocol and critical review of the manuscript: Yaël Pinhas, Jérémie F. Cohen, Rachel Y. Moon, Floortje Kanits, Sabine Plancoulaine, Anne-Laure Sellier, Inge Harrewijn, Christèle Gras-Le Guen

Final approval of the version to be published: all authors

## Data Availability

All data produced in the present study are available upon reasonable request to the authors

## ONLINE-ONLY SUPPLEMENTS

**eTable 1** PRISMA 2020 Checklist

**eTable 2** Risk of bias assessment items (adapted from Hoy *et al.*, J Clin Epidemiol 2012)

**eTable 3** Risk of bias assessment results

**eText** Search box in PubMed

