## Supplements for "Inconsistency of infant caretakers’ visual exposome with safe infant sleep recommendations: a systematic review and meta-analysis"

**ONLINE-ONLY SUPPLEMENTS**

**eTable 1** PRISMA 2020 Checklist

**eTable 2** Risk of bias assessment items (adapted from Hoy *et al.*, J Clin Epidemiol 2012)

**eTable 3** Risk of bias assessment results

**eText** Search box in PubMed

**eTable 1** PRISMA 2020 Checklist

| Section and Topic | Item # | Checklist item | Location where item is reported |
|---|---|---|---|
| **TITLE** | | | |
| Title | 1 | Identify the report as a systematic review. | 1 |
| **ABSTRACT** | | | |
| Abstract | 2 | See the PRISMA 2020 for Abstracts checklist. | 2 |
| **INTRODUCTION** | | | |
| Rationale | 3 | Describe the rationale for the review in the context of existing knowledge. | 3-4 |
| Objectives | 4 | Provide an explicit statement of the objective(s) or question(s) the review addresses. | 4 |
| **METHODS** | | | |
| Eligibility criteria | 5 | Specify the inclusion and exclusion criteria for the review and how studies were grouped for the syntheses. | 4 |
| Information sources | 6 | Specify all databases, registers, websites, organisations, reference lists and other sources searched or consulted to identify studies. Specify the date when each source was last searched or consulted. | 4 |
| Search strategy | 7 | Present the full search strategies for all databases, registers and websites, including any filters and limits used. | 4-5 |
| Selection process | 8 | Specify the methods used to decide whether a study met the inclusion criteria of the review, including how many reviewers screened each record and each report retrieved, whether they worked independently, and if applicable, details of automation tools used in the process. | 5 |
| Data collection process | 9 | Specify the methods used to collect data from reports, including how many reviewers collected data from each report, whether they worked independently, any processes for obtaining or confirming data from study investigators, and if applicable, details of automation tools used in the process. | 5-6 |
| Data items | 10a | List and define all outcomes for which data were sought. Specify whether all results that were compatible with each outcome domain in each study were sought (e.g. for all measures, time points, analyses), and if not, the methods used to decide which results to collect. | 5 |
| | 10b | List and define all other variables for which data were sought (e.g. participant and intervention characteristics, funding sources). Describe any assumptions made about any missing or unclear information. | 6 |
| Study risk of bias assessment | 11 | Specify the methods used to assess risk of bias in the included studies, including details of the tool(s) used, how many reviewers assessed each study and whether they worked independently, and if applicable, details of automation tools used in the process. | 5-6 |
| Effect measures | 12 | Specify for each outcome the effect measure(s) (e.g. risk ratio, mean difference) used in the synthesis or presentation of results. | 6 |
| Synthesis methods | 13a | Describe the processes used to decide which studies were eligible for each synthesis (e.g. tabulating the study intervention characteristics and comparing against the planned groups for each synthesis (item #5)). | 5-6 |
| | 13b | Describe any methods required to prepare the data for presentation or synthesis, such as handling of missing summary statistics, or data conversions. | n/a |
| | 13c | Describe any methods used to tabulate or visually display results of individual studies and syntheses. | 6 |

| Section and Topic | Item # | Checklist item | Location where item is reported |
|---|---|---|---|
| | 13d | Describe any methods used to synthesize results and provide a rationale for the choice(s). If meta-analysis was performed, describe the model(s), method(s) to identify the presence and extent of statistical heterogeneity, and software package(s) used. | 6 |
| | 13e | Describe any methods used to explore possible causes of heterogeneity among study results (e.g. subgroup analysis, meta-regression). | n/a |
| | 13f | Describe any sensitivity analyses conducted to assess robustness of the synthesized results. | n/a |
| Reporting bias assessment | 14 | Describe any methods used to assess risk of bias due to missing results in a synthesis (arising from reporting biases). | 6-7 |
| Certainty assessment | 15 | Describe any methods used to assess certainty (or confidence) in the body of evidence for an outcome. | 6-7 |
| **RESULTS** | | | |
| Study selection | 16a | Describe the results of the search and selection process, from the number of records identified in the search to the number of studies included in the review, ideally using a flow diagram. | 7 |
| | 16b | Cite studies that might appear to meet the inclusion criteria, but which were excluded, and explain why they were excluded. | 7 |
| Study characteristics | 17 | Cite each included study and present its characteristics. | 7 |
| Risk of bias in studies | 18 | Present assessments of risk of bias for each included study. | 7-8 |
| Results of individual studies | 19 | For all outcomes, present, for each study: (a) summary statistics for each group (where appropriate) and (b) an effect estimate and its precision (e.g. confidence/credible interval), ideally using structured tables or plots. | 8 |
| Results of syntheses | 20a | For each synthesis, briefly summarise the characteristics and risk of bias among contributing studies. | 8 |
| | 20b | Present results of all statistical syntheses conducted. If meta-analysis was done, present for each the summary estimate and its precision (e.g. confidence/credible interval) and measures of statistical heterogeneity. If comparing groups, describe the direction of the effect. | 8 |
| | 20c | Present results of all investigations of possible causes of heterogeneity among study results. | n/a |
| | 20d | Present results of all sensitivity analyses conducted to assess the robustness of the synthesized results. | n/a |
| Reporting biases | 21 | Present assessments of risk of bias due to missing results (arising from reporting biases) for each synthesis assessed. | 10 |
| Certainty of evidence | 22 | Present assessments of certainty (or confidence) in the body of evidence for each outcome assessed. | 11 |
| **DISCUSSION** | | | |
| Discussion | 23a | Provide a general interpretation of the results in the context of other evidence. | 9-10 |
| | 23b | Discuss any limitations of the evidence included in the review. | 9-10 |
| | 23c | Discuss any limitations of the review processes used. | 9-10 |
| | 23d | Discuss implications of the results for practice, policy, and future research. | 10-11 |

| Section and Topic | Item # | Checklist item | Location where item is reported |
|---|---|---|---|
| **OTHER INFORMATION** | | | |
| Registration and protocol | 24a | Provide registration information for the review, including register name and registration number, or state that the review was not registered. | n/a |
| | 24b | Indicate where the review protocol can be accessed, or state that a protocol was not prepared. | n/a |
| | 24c | Describe and explain any amendments to information provided at registration or in the protocol. | n/a |
| Support | 25 | Describe sources of financial or non-financial support for the review, and the role of the funders or sponsors in the review. | 12-13 |
| Competing interests | 26 | Declare any competing interests of review authors. | 12-13 |
| Availability of data, code and other materials | 27 | Report which of the following are publicly available and where they can be found: template data collection forms; data extracted from included studies; data used for all analyses; analytic code; any other materials used in the review. | 12-13 |

**eTable 2** Risk of bias assessment items (adapted from Hoy *et al.*, J Clin Epidemiol 2012)

| **Risk of bias items** | **Signaling question** | **Evaluation criteria and examples** | **Responses** | |
|---|---|---|---|---|
| | | | **Low risk** | **High risk** |
| **Internal validity** | | | | |
| 1. Directly collected | Were data collected directly from the subjects (as opposed to a proxy)? | Data were collected directly from the pictures evaluated by experts, no proxy use | Yes | No/unclear |
| 2. Acceptable case definition | Was an acceptable case definition used in the study? | List recommendations individually or refer to those of public health agencies or regulatory or academic societies in pediatrics | Yes | No/unclear |
| 3. Reliable and valid evaluation | Was the study instrument that measured the parameter shown to have validity and reliability? | Independent evaluation by several experts and adequate management of disagreements | Yes | No/unclear |
| 4. Same mode of data collection | Was the same mode of data collection used for all subjects? | Same criteria evaluated for all pictures | Yes | No/unclear |
| 5.Length of the shortest prevalence period | Was the length of the shortest prevalence period for the parameter of interest appropriate? | Not relevant | | |
| 6. Appropriate numerator(s) and denominator(s) for the parameter of interest | Were the numerator(s) and denominator(s) for the parameter of interest appropriate? | Numerator and denominator clearly defined | Yes | No/unclear |
| **External validity** | | | | |
| 7.Representative of national population | Was the study's target population close representation of the national population in relation to relevant variables? | Not relevant | | |
| 8. Representative of target population | Was the sampling frame a true or close representation of the target population? | Restrictions were applied to studied visual public spaces, e.g. the first 100 search results, female readership, … | No | Yes/unclear |
| 9. Random selection | Was some form of random selection used to select the sample, OR was a census undertaken? | Random selection by keyword search or census of a type of visual public space during a specific period, … | Yes | No/unclear |
| 10. Non-response bias | Was the likelihood of nonresponse bias minimal? | Not relevant | | |

**eTable 3** Risk of bias assessment results

| Author (year), by date of publication | Internal validity | | | | External validity | | | Risk of bias score** | |
|---|---|---|---|---|---|---|---|---|---|
| | Directly collected | Acceptable case definition | Reliable and valid evaluation | Same mode of data collection | Representative of target population | Random selection | Appropriate numerator(s) and denominator(s) for the parameter of interest | Overall score | Overall risk of bias |
| Joyner *et al.* (2009) | 0 | 0 | 0 | 0 | 1 | 0 | 0 | 1 | low |
| Epstein *et al.* (2011) | 0 | 0 | 0 | 1 | 1 | 0 | 0 | 2 | low |
| Goodstein *et al.* (2018) | 0 | 0 | 0 | 0 | 1 | 0 | 0 | 1 | low |
| Kamra *et al.* (2018) | 0 | 0 | 0 | 0 | 1 | 0 | 0 | 1 | low |
| Chin *et al.* (2021) | 0 | 0 | 0 | 0 | 1 | 0 | 0 | 1 | low |
| Kanits *et al.* (2023) | 0 | 0 | 0 | 0 | 1 | 0 | 0 | 1 | low |
| De Visme *et al.* (2023) | 0 | 0 | 0 | 0 | 1 | 0 | 0 | 1 | low |

* risk: 0=low risk; 1=high risk
** Risk of bias score from Hoy *et al*, J Clin Epidemiol 2012 and Brennan *et al*, BMJ Open 2024: low (0–2), moderate (3–5) and high (6–7) risk

**eText** Search box in PubMed

(infant* or baby or babies or newborn*) AND (sleep*) AND (safe* or unsafe or environment*) AND (recommendation* or guid* or advi*)
